# Muscle strength and muscle mass in people with long term conditions and multimorbidity: A cross-sectional study of UK Biobank participants

**DOI:** 10.1101/2025.08.11.25333208

**Authors:** Marion Guerrero-Wyss, Stuart Johnston, Fanny Peterman-Rocha, Irene Rodríguez-Gómez, Lyn D Ferguson, Frederick K Ho, Bhautesh D Jani, Stuart R Gray, Carlos A Celis-Morales

## Abstract

**BACKGROUND/OBJECTIVES:** To compare muscle strength and muscle mass in individuals with various long-term conditions (LTCs) to those of healthy individuals, using data from the UK Biobank.

**SUBJECTS/METHODS:** This cross-sectional study of 444,420 UK Biobank participants (aged 37 – 73) examined 25 self-reported LTCs ( ≥ 1% prevalence). Multimorbidity was defined as a prevalent LTCs count ≥ 2. Muscle strength was assessed using a handgrip dynamometer, while muscle mass (fat-free mass as a percentage of body weight) was assessed using BIA, DXA, and MRI; all expressed as a percentage of total body weight. Linear regression analyses were conducted to examine how muscle strength and muscle mass varied by LTCs and multimorbidity.

**RESULTS:** The lowest muscle strength was observed in men with rheumatoid arthritis (RA), type 2 diabetes (T2D), stroke, gout, and deep venous thrombosis (-11.8% to -6.1%) lower than the reference group and in women with RA, gout, T2D, coronary heart disease (CHD), and stroke (-11.8% to -6.5%). Muscle mass was lowest in men with T2D, gout, CHD, hypertension, and high cholesterol (-4.7% to -3.1%), and in women with gout, T2D, hiatus hernia, CHD, and hypertension (-6.4% to -4.1%). Trends were consistent across measurements using DXA (n=4,719) and MRI (n=22,901). Muscle strength and muscle mass were lower in people with more LTCs, with a dose-response relationship.

**CONCLUSIONS:** This study demonstrates that muscle strength and muscle mass were lower in people with multiple LTCs and specific conditions. Emphasizing the need for targeted muscle interventions, particularly for those with T2D, RA, gout, and CHD.

## INTRODUCTION

Maintaining adequate levels of muscle strength and muscle mass is crucial for maintaining good health, particularly in middle-aged and older adults (1, 2). A growing body of evidence indicates that low levels of muscle strength and muscle mass are associated with an increased risk of falls, fractures, and long-term conditions (LTCs) such as cardiovascular diseases, dementia, certain cancers, and even mortality (1–5).

Evidence shows that grip strength predicts cardiovascular and all-cause mortality and declines as the number of LTCs increases (2). Additionally, other studies have found that muscle strength is progressively lower with an increasing number of LTCs an individual has

(3). This is important as the number of people living with two or more LTCs, termed multimorbidity, is increasing, and especially in younger individuals and deprived areas (6, 7); and has become a growing priority for health systems and global health research efforts.

Several limitations in previous studies should be noted, including small sample sizes, a lack of adjustment, and the reliance on grip strength as the sole measure of muscle strength without using other assessment methods (1, 3, 8).

The impact of long-term conditions (LTCs) on muscle mass is not well understood, nor is there a comprehensive understanding of which specific conditions have the most detrimental impact on muscle strength and mass. Filling these gaps could inform more tailored strength training guidelines for individuals with LTCs. This study aims to compare muscle strength and muscle mass in individuals with various LTCs, to those with no LTCs; and by LTC count using data from the UK Biobank.

## METHODS

### Study design and participants

From 2006 to 2010, UK Biobank recruited over 500,000 participants (5.5% response rate), aged 37 to 73 years old from the general population (9). Participants visited one out of 22 assessment centres across England, Wales, and Scotland, and completed a touch-screen questionnaire, had anthropometry, body composition (assessed by bio-impedance (BIA)) and grip strength assessed, and biological samples taken, as described in detail elsewhere (10, 11). In 2014 the UK Biobank imaging enhancement protocol was initiated incorporating full body Magnetic Resonance Imaging (MRI) and X-ray absorptiometry (DXA) scans. The UK Biobank has amassed one of the largest datasets utilizing objective methods for quantifying strength levels as well as muscle mass using different assessment methods including DXA, MRI, and BIA.

### Ethical approval

The UK Biobank study was approved by the Northwest Multicentre Research Ethics Committee and performed in accordance with the ethical standards laid down in the 1964 Declaration of Helsinki and its later amendments. Informed consent was given by all participants at the time of recruitment. This study is part of the UK Biobank project 7155 (NHS National Research Ethics Service16/NW/0274).

### Outcomes assessment Muscle strength

Hand grip strength was used as a proxy of muscle strength (3), and measured using a Jamar J00105 hydraulic hand dynamometer (12). Participants sat upright with their elbows on the side and bent 90 degrees so that their forearms were facing forward and resting on the armrest (13). Maximum grip strength of the right and left arms over three seconds was recorded (13).

The maximum value (in kg) for either arm was used for analysis (12).

### Muscle mass

Fat-free mass (FFM), a marker of muscle mass, was assessed by BIA, DXA and MRI and expressed as kg or liters of FFM. BIA was performed with Tanita BC418MA Body Composition Analyser (Tanita, Japan). The analyser measured body impedance with a high-frequency current (50 kHz) and 8 contact electrodes. Participants were asked to place their bare feet on the analyser platform, to keep their feet still and in good contact with the platform’s metallic electrodes on platform, and to grip the two handles firmly with palm and thumb contacting metallic electrodes and arms hanging loosely by their sides.

DXA was performed with Lunar iDXA Scanner (General Electric Healthcare, Wisconsin US). The scanner was calibrated daily following standard quality control procedures to maintain consistency. Participants were asked to lie flat on their back on the scanning couch for whole body scan. All operators had standardised central training, and all scans were performed according to standard operating procedures.

For the MRI assessment participants were scanned in a Siemens MAGNETOM Aera 1.5-T MRI scanner (Siemens Healthineers, Erlangen, Germany) using a 6 min dual-echo Dixon Vibe protocol, providing a water and fat separated volumetric data set covering neck to knees. Body composition analyses were performed using AMRA® Researcher (AMRA Medical AB, Linköping, Sweden (14, 15). Fat-tissue free muscle volume was defined as the volume of all voxels with fat fraction <50% (‘viable muscle tissue’) in the thighs. This is to be differentiated from lean muscle volume (sometimes called ‘contractile muscle volume’) calculated by subtracting the fat volume from the entire muscle volume. Detailed muscle composition measurements and definitions have been previously described in previous studies (15). All muscle strength and mass measurements were presented as a percentage of body weight to account for variations in body size and to facilitate comparisons across the assessments used to estimate FFM.

### Long Term Conditions and multimorbidity

Self-reported long-term conditions (LTCs) were identified through a nurse-led questionnaire at baseline, where participants reported any serious illnesses or disabilities diagnosed by a doctor. LTCs were classified using the International Classification of Diseases, Tenth Revision (ICD-10) (16). There were 322 LTCs coded in the UK Biobank. Of these, 25 self-reported LTCs with a prevalence >1% were included in the current study. The LTCs included were hypertension, high cholesterol, asthma, osteoarthritis, hiatal hernia, depression, type 2 diabetes (T2D), allergic rhinitis, hypothyroidism, coronary heart diseases (CHD), migraine, eczema, breast cancer (for women), prostate cancer (for men), deep venous thrombosis (DVT), anxiety, stroke, back problems, chronic obstructive pulmonary diseases (COPD), osteoporosis, gout, emphysema, rheumatoid arthritis (RA), psoriasis, diverticulitis and glaucoma, as presented in Supplementary Table 1. Participants without any of the 322 LTCs were categorized as healthy and served as the reference group (n=173,414).

Multimorbidity was defined as having two or more LTCs. LTCs were examined both as individual LTCs, and collectively as a measure of multimorbidity, by counting LTCs and categorising into: 0, 1, 2, 3, 4 or ≥5 LTCs.

### Covariates

These included age, Townsend deprivation index, alcohol intake, smoking and ethnicity. Age was calculated based on dates of birth and baseline assessment. The Townsend deprivation index was derived from the postcode of residence. Ethnicity was defined in five categories: White, Mixed, South Asian, Black and Chinese. Smoking status was categorized in never, previous and current. Alcohol intake was self-reported and combined as units per week.

### Statistical analyses

Descriptive cohort characteristics by categories of LTCs are presented as mean and standard deviation (SD) for quantitative variables and as a proportion for categorical variables. Data were normally distributed, and Pearson Correlation was performed to compare the relationship between grip strength and muscle mass outcomes assessed with BIA, DXA and MRI.

Linear regression analyses were conducted to examine the association between individuals’ LTCs and markers of muscle strength and muscle quantity. These markers included strength assessed with a hand dynamometer, and muscle mass assessed via BIA, DXA, and MRI. Results were reported as means along with their 95% confidence intervals (CIs). Similar analyses were conducted to explore the association of the number LTCs, categorized as 0, 1, 2, 3, 4, and 5+ LTCs, with muscle strength and muscle mass. All analyses were stratified by sex and adjusted for age, deprivation index, ethnicity, smoking status, and alcohol consumption.

To account for potentially inflated type I errors due to multiple testing, all analyses were adjusted using Holm’s method (P_adj_) (17, 18), and the significance threshold was set at p < 0.0001. Analyses were conducted using STATA 18 (StataCorp LP).

## RESULTS

Of the 502,350 participants recruited to the UK Biobank (53.8% women, 46.2% men), data on muscle strength and muscle mass were available for 444,420 participants measured by BIA, 4,653 by DXA, 22,091 by MRI, and 444,420 for grip strength (Supplementary Figure 1 and Supplementary Table 2).

The characteristics of the cohort, stratified by the number of long-term conditions (LTCs), are summarized in Supplementary Table 3. In total, 34.8% of participants reported having no LTCs, 32.9% reported one LTC, and 32.3% reported two or more LTCs. Participants with multimorbidity tended to be older, were more likely to be women, had higher body mass index (BMI), engaged in lower levels of physical activity, had lower muscle strength and fat-free mass (FFM), and were more likely to have obesity and be former smokers compared to those without LTCs. The differences were even more pronounced for individuals with five or more LTCs (Supplementary Table 3).

The correlations between the outcomes, including muscle strength and FFM are presented in Supplementary Figure 2. The correlations were strong, ranging from 0.692 to 0.924. The weakest correlation was observed between muscle strength and FFM measured by DXA (r = 0.692), while the strongest correlation was between FFM measured by MRI and DXA (r = 0.924).

Figures 1-4 illustrate muscle strength and muscle mass across the 25 most prevalent LTCs. For handgrip strength, the five LTCs associated with the lowest strength in men were RA (-11.6%), T2D (-9.07 %), stroke (-6.78%), gout (-6.49 kg), and DVT (-6.09 %), compared to those without LTCs (reference group) (Figure 1). In women, the five conditions with the lowest strength were RA (-11.8%), gout (-9.05%), T2D (-8.72%), CHD (-7.07%), and stroke (-6.57%), compared to the reference group (Figure 1). Absolute handgrip strength values, expressed in kilograms, for each of the 25 LTCs are presented in Figure 1.

**Figure 1.**
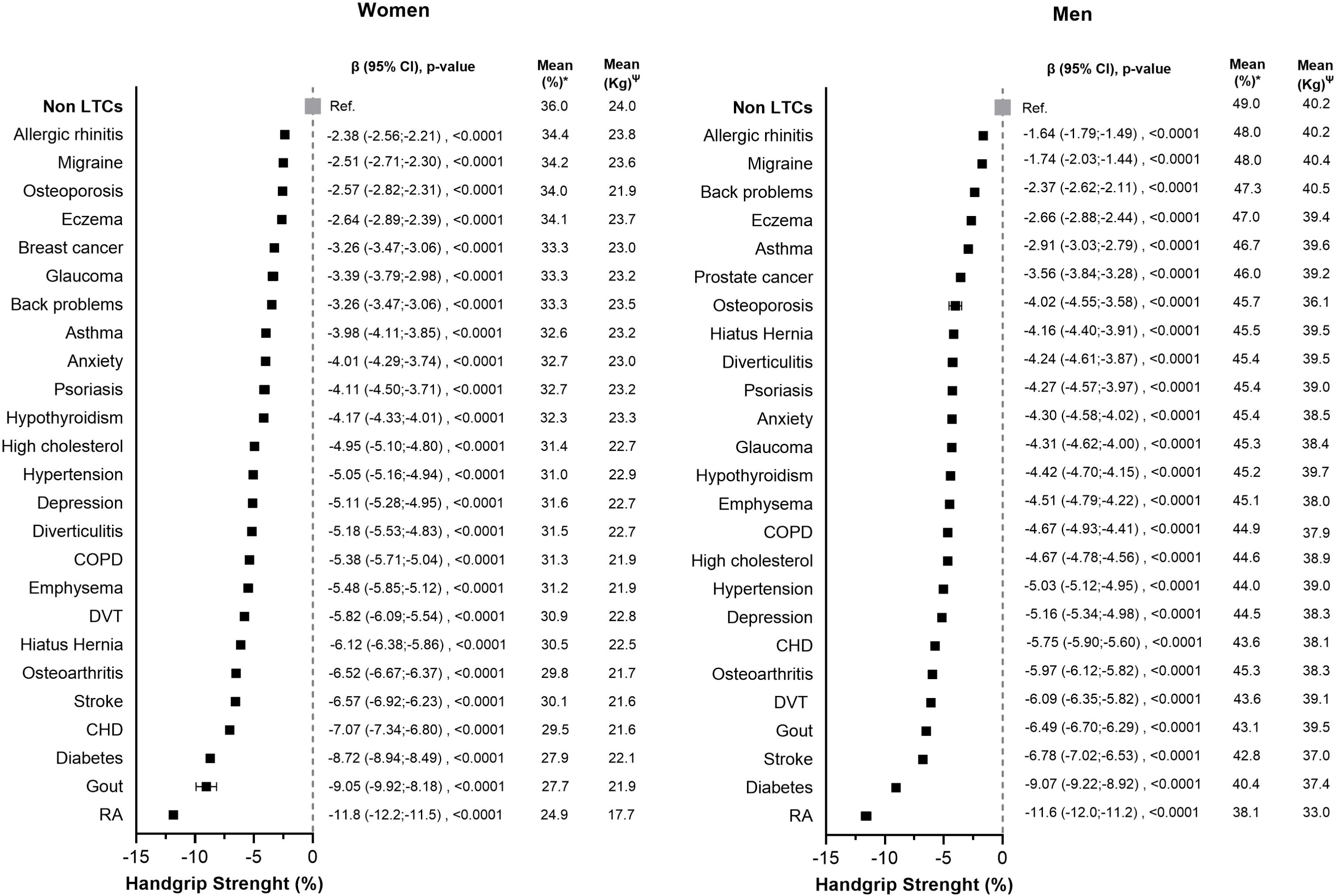
Differences in Strength Levels by Long-Term Conditions in women and men. The forest plot displays the differences in handgrip strength, expressed as a percentage of body weight, expressed as a percentage of body weight, along with their 95% confidence intervals, in individuals with LTCs compared to those without (reference group). Muscle strength means are presented both as a percentage of body weight (*) and in liters (Ψ absolute values). The analyses were adjusted for age, smoking status, alcohol intake, deprivation index, and ethnicity. Abbreviations: Kg, kilograms; CI, confidence intervals; COPD, chronic obstructive pulmonary disease; CHD, coronary heart disease; DVT, deep vein thrombosis; RA, rheumatoid arthritis.

Muscle mass, estimated by BIA, was lower in men with T2D (-4.71%), gout (-4.25%), CHD (-3.50%), hypertension (-3.24%), and high cholesterol (-3.15%), in comparison to the reference group (Figure 2). In women the lowest muscle mass was seen in gout (-6.42%), T2D (-5.89%), hiatus hernia (-4.68%), CHD (-4.35%), and hypertension (-4.14%), compared to the reference group (Figure 2). Muscle mass, measured by DXA, showed slightly different patterns: for men, the top five LTCs with the lowest muscle mass were type 2 diabetes (- 6.08%), RA (-5.65%), hiatus hernia (-3.80%), gout (-3.63%), and COPD (-3.55%), lower than the reference group (Figure 3). In women, hiatus hernia (-5.74%), T2D (-5.10%), COPD (-4.22%), CHD (-4.16%), and emphysema (-3.77%) had the lowest muscle mass, compared to the reference group (Figure 3). Thigh muscle mass, measured by MRI, revealed similar trends, with the lowest levels in men seen in T2D (-1.88%), gout (-1.11%), CHD (-0.99%), diverticulitis (-0.92%), and hypertension (-0.92%), compared to the reference group (Figure 4). In women, T2D (-1.51%), COPD (-1.15%), hiatus hernia (-1.09%), gout (-1.04%), and emphysema (-0.98%) were associated with the lowest levels, in comparison to the reference group (Figure 4). Absolute muscle mass values, expressed in liters or Kg, for each of the 25 LTCs are presented in Figure 2-4.

**Figure 2.**
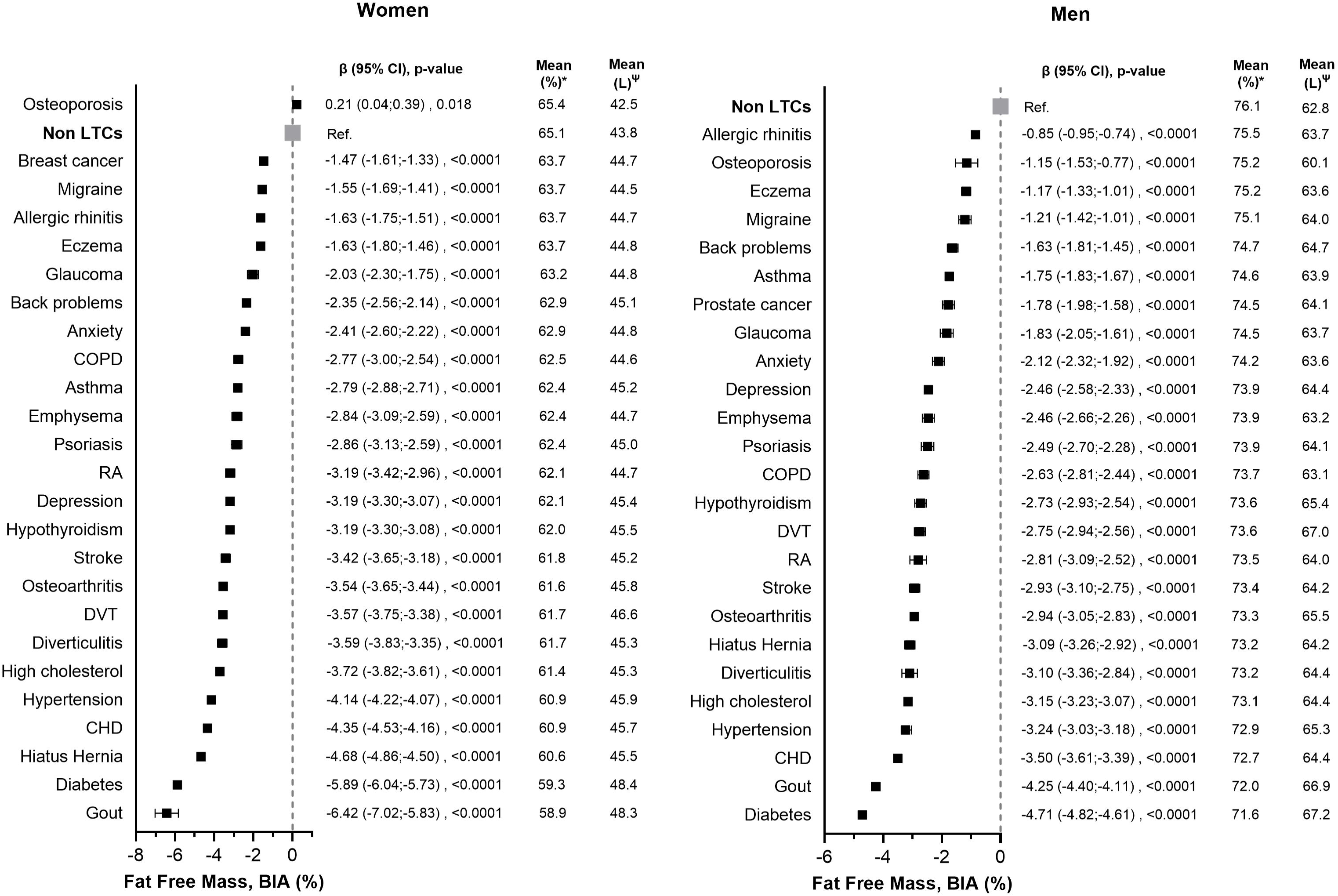
Differences in BIA Fat Free Mass Levels by Long-Term Conditions in women and men. The forest plot illustrates the differences in Fat-Free Mass (FFM) measured by BIA, expressed as a percentage of body weight, along with their 95% confidence intervals, in individuals with LTCs compared to those without (reference group). FFM means are presented both as a percentage of body weight (*) and in liters (Ψ absolute values). The analyses were adjusted for age, smoking status, alcohol intake, deprivation index, and ethnicity. Abbreviations: Kg, kilograms; CI, confidence intervals; COPD, chronic obstructive pulmonary disease; CHD, coronary heart disease; DVT, deep vein thrombosis; RA, rheumatoid arthritis.

**Figure 3.**
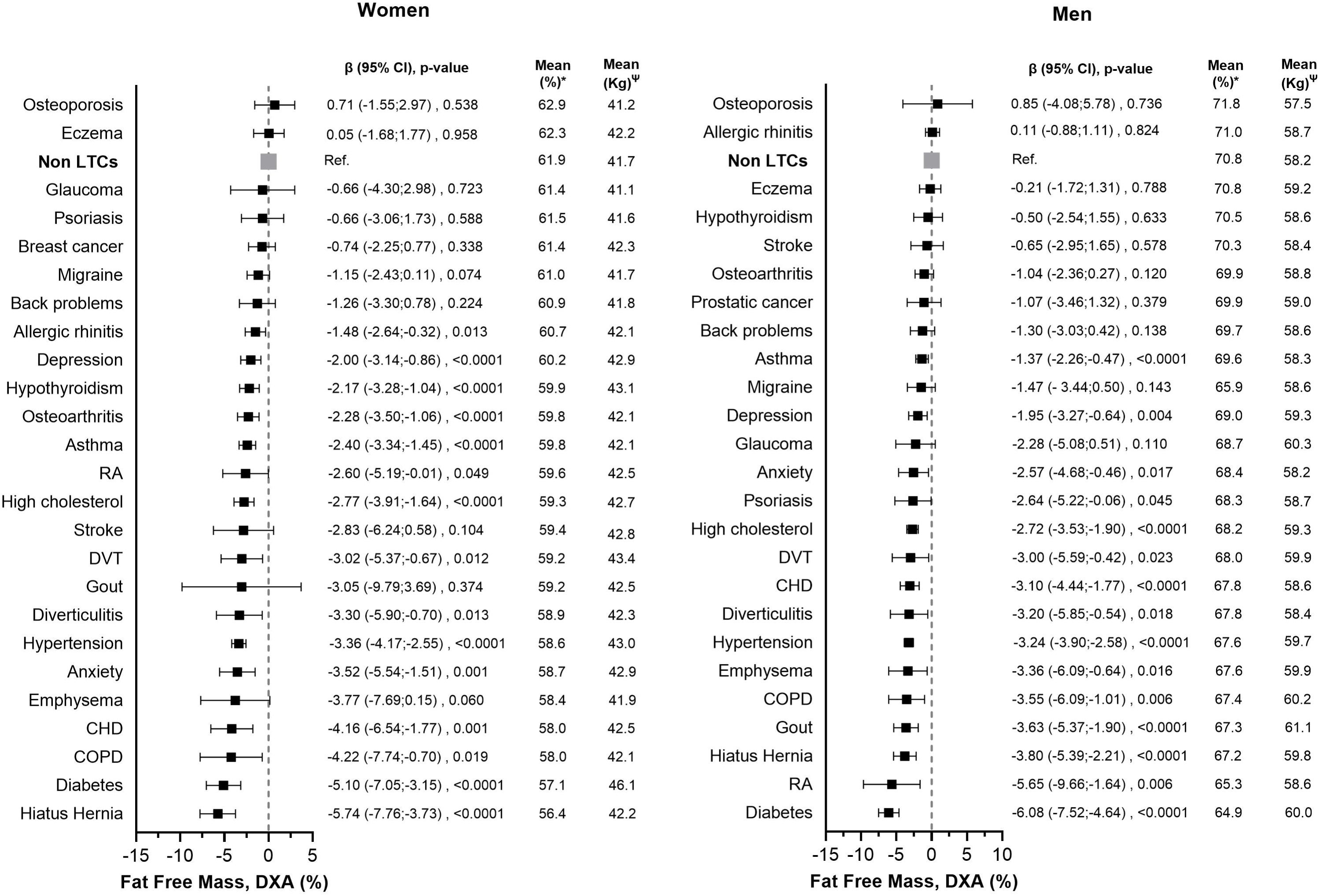
Differences in DXA Fat Free Mass Levels by Long-Term Conditions in women and men. The forest plot illustrates the differences in Fat-Free Mass (FFM) measured by DXA, expressed as a percentage of body weight, along with their 95% confidence intervals, in individuals with LTCs compared to those without (reference group). FFM means are presented both as a percentage of body weight (*) and in liters (Ψ absolute values). The analyses were adjusted for age, smoking status, alcohol intake, deprivation index, and ethnicity. Abbreviations: Kg, kilograms; CI, confidence intervals; COPD, chronic obstructive pulmonary disease; CHD, coronary heart disease; DVT, deep vein thrombosis; RA, rheumatoid arthritis.

**Figure 4.**
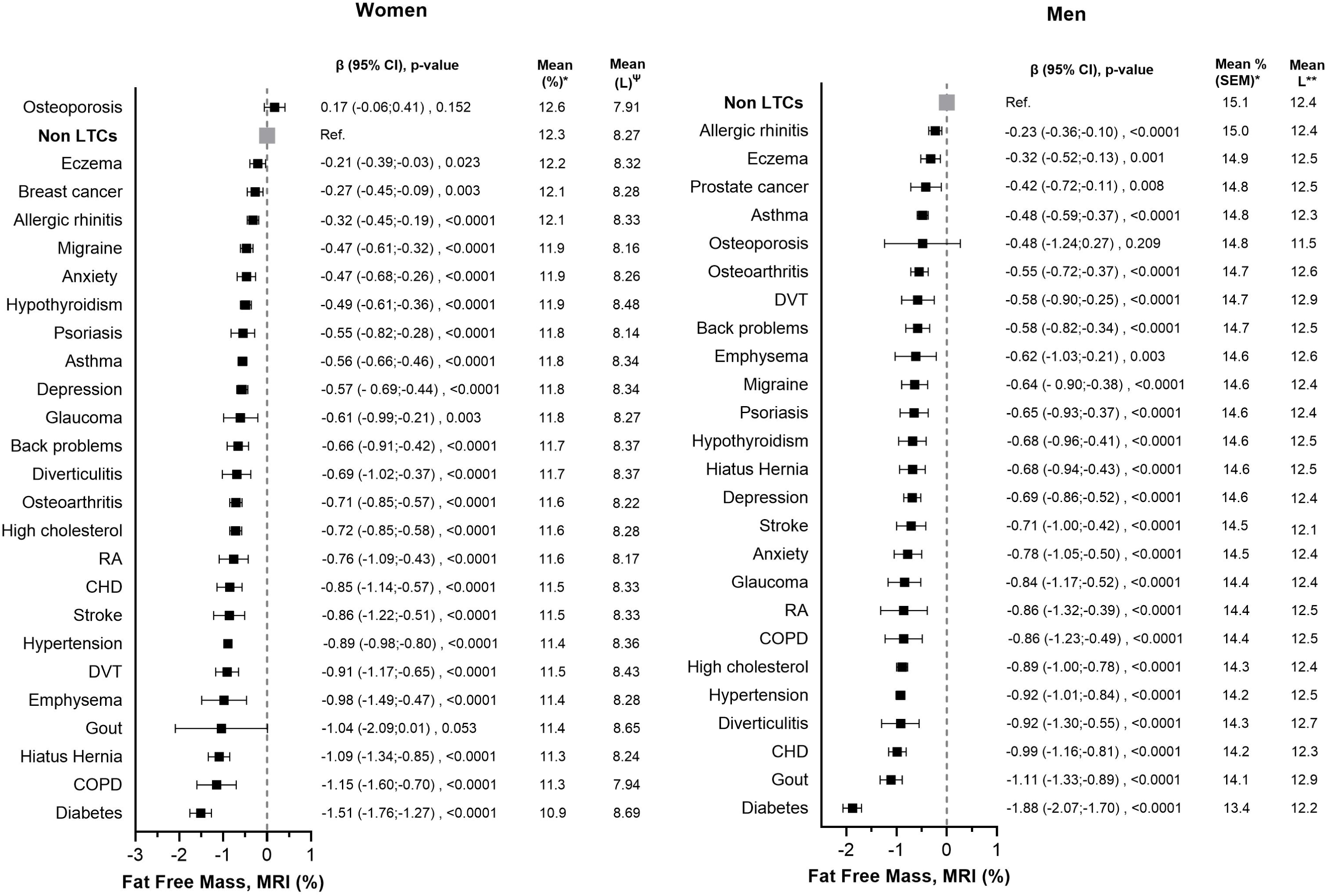
Differences in MRI Fat Free Mass Levels by Long-Term Conditions in women and men. The forest plot illustrates the differences in Fat-Free Mass (FFM) measured by MRI, expressed as a percentage of body weight, along with their 95% confidence intervals, in individuals with LTCs compared to those without (reference group). FFM means are presented both as a percentage of body weight (*) and in liters (Ψ absolute values). The analyses were adjusted for age, smoking status, alcohol intake, deprivation index, and ethnicity. Abbreviations: Kg, kilograms; CI, confidence intervals; COPD, chronic obstructive pulmonary disease; CHD, coronary heart disease; DVT, deep vein thrombosis; RA, rheumatoid arthritis.

Figure 5 presents the associations between LTC count and muscle strength and muscle mass. Both muscle strength and muscle mass showed an inverse dose-response relationship with LTC count. For each additional LTC, muscle strength was lower on average by 1.81% in men and 1.71% in women. Strength levels were 10.6% lower in men and 9.96% lower in women with 5 or more LTCs compared to those without LTCs. FFM, assessed by BIA, was 0.94% lower in men and 1.03% lower in women per additional LTC. In individuals with 5 or more LTCs, FFM levels were 5.14% lower in men and 5.66% lower in women compared to those without LTCs. Comparable trends were observed for FFM, assessed by DXA and MRI, as shown in Figure 5. Levels of muscle strength and muscle mass expressed in absolute terms (kg or liters) by number of LTCs are presented in Supplementary Figure 3.

**Figure 5.**
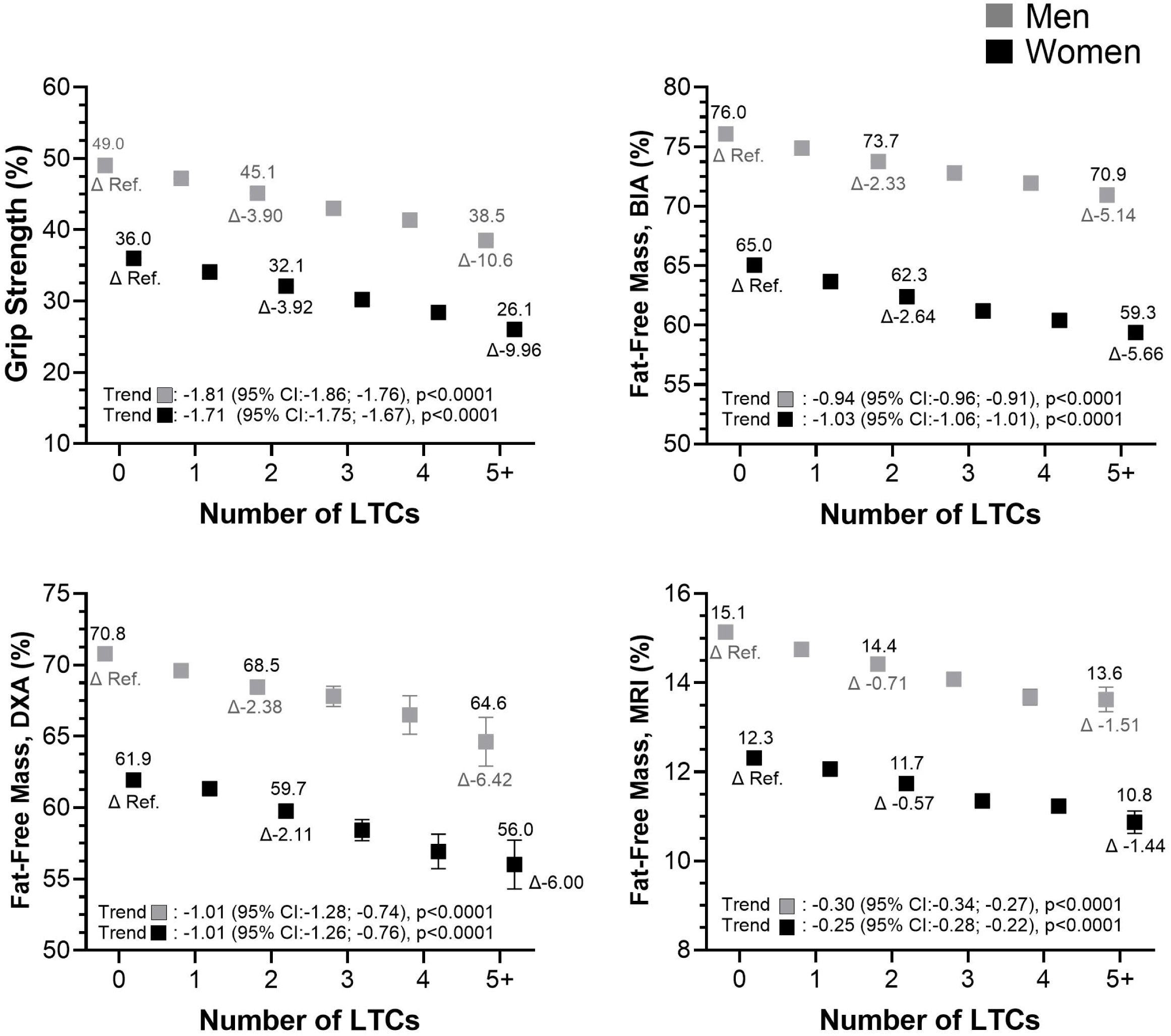
Levels of strength and Fat-free mass by number of LTCs in men and women. The data present adjusted means and their 95% confidence intervals (CI) for handgrip strength and Fat-Free Mass (FFM) levels, assessed via BIA, DXA, and MRI, all reported as a percentage of body weight. Differences between individuals with no LTCs, 2 LTCs, and 5 or more LTCs are displayed, with means shown above the plotted squares and the deltas presented below (Δ). Trend values indicate the mean change in the outcome per 1 LTC increment. The analyses were adjusted for age, ethnicity, deprivation, smoking status, and alcohol intake. Kg, kilograms; CI, confidence intervals.

## DISCUSSION

This study demonstrates that there is significantly lower muscle strength and mass, regardless of measurement method, in people with the 25 most prevalent LTCs compared with people without any LTCs. Our results also show an inverse dose-response relationship between LTC count and both muscle strength and mass. Specifically, individuals with five or more LTCs had about 10% lower strength and 5-6% lower muscle mass compared to those without any LTCs. Of all the included LTCs, people with RA had the lowest muscle strength, and people with T2D had the lowest muscle mass. These findings highlight the importance of directing the limited resource, to the clinical population where it could have the most benefits, and the greatest impact, aimed at increasing muscle strength and mass, particularly for individuals with T2D, RA, or those with multimorbidity.

While this study does not establish causality, the strong association between LTCs and lower muscle strength and muscle mass in both sexes may stem from common mechanisms, particularly inflammation and low physical activity. Cardiovascular diseases (e.g., stroke, hypertension, CHD), metabolic disorders (e.g., T2D), and inflammatory arthritides (e.g., RA and gout) trigger inflammatory pathways that impair muscle protein synthesis, leading to muscle atrophy and weakness (19, 20). Indeed, elevated cytokines, such as IL-6 and TNF-α, are linked to muscle loss in both healthy and diseased individuals (21, 22) and chronic inflammation contributes to skeletal muscle mass loss (23) and muscle degradation (24). Physical activity helps mitigate these effects by promoting anabolism, reducing catabolism, and decreasing inflammation, which may explain the lower muscle strength and mass in LTC populations with lower levels of physical activity (12, 25).

In our study, musculoskeletal diseases, particularly rheumatoid arthritis, were associated with the lowest levels of handgrip strength, likely due to inflammation, joint damage, and resulting physical disability. Similar findings have been reported in individuals with RA, highlighting significant declines in muscle mass and strength, even among patients with early-stage RA (26). In another study, RA patients had 10% less appendicular lean mass, and a 27% higher body fat percentage compared to healthy controls, with muscle function measures being 24- 34% poorer in RA patients (27). Additionally, muscle tissue does not fully recover, even during periods of clinical remission, suggesting that rheumatoid arthritis may lead to persistent, long-term muscle damage (26).

These conditions lead to disuse atrophy and contractile dysfunction (28). RA, in particular, is characterized by progressive muscle loss, with sarcopenia masked by increased adiposity, known as ’rheumatoid cachexia.’ Recent studies highlight a higher prevalence of sarcopenic obesity in rheumatoid arthritis compared to non-RA controls, which is linked to functional decline and increased disability over time (29, 30). This underscores the importance of muscle-strengthening interventions, such as resistance exercise, in this population, especially considering the high multimorbidity prevalence (31). In our study, RA was most strongly associated with low grip strength and reduced muscle mass, placing this population at significantly higher risk. Low levels of strength and muscle mass are linked to an increased risk of both all-cause and cause-specific mortality (32), exponentially elevating the risk for these patients. These findings underscore the importance of implementing appropriate strategies to improve muscle health in individuals with RA.

Metabolic and cardiovascular conditions, including T2D, stroke, and CHD, are also strongly associated with lower muscle mass and strength (8, 33). Notably, higher levels of handgrip strength have been linked to significantly reduced event rates of cardiovascular disease (CVD) (34). In fact, handgrip strength has been recognized as a predictor of CVD incidence (8, 34).

Other conditions, such as hiatal hernia, though less frequently discussed, also contribute to reduced muscle mass due to physical inactivity and nutritional challenges (35, 36). In older adults, the combination of sarcopenia and hiatal hernia can exacerbate muscle loss (37). Additionally, obesity is a major risk factor for hiatal hernia, and individuals with multimorbidity generally have higher BMI and lower muscle mass compared to healthy participants (38).

Although muscle mass measurements assessed by BIA, DXA, and MRI show moderate to strong correlations, notable differences exist among these muscle mass phenotypes. The discrepancies in fat-free mass (FFM) estimates obtained by DXA, bioimpedance, and MRI in individuals with long-term conditions (LTCs) could be attributed to variations in measurement techniques, the influence of LTCs on body composition, and the scope of each method. Fat-free or lean tissue, as assessed by DXA and bioimpedance, encompasses not only skeletal muscle but also organs, bone, bodily fluids, and water content. In contrast, MRI provides a more precise quantification of true muscle tissue, particularly in the thigh region.

Additionally, MRI offers valuable insights into muscle quality by assessing the extent of intramuscular fat infiltration, often revealing more pronounced muscle loss in specific areas (15, 39). Additionally, LTCs may alter tissue composition, affecting the accuracy of these methods in estimating true muscle mass. These factors together may explain the discrepancies in FFM measurements across different techniques in this population.

This study highlights the critical importance of prioritizing muscle health in individuals with LTCs. It reveals significantly lower muscle strength and mass among those with the 25 most prevalent LTCs compared to healthy individuals, with a dose-response relationship showing further declines as the number of LTCs increases. Notably, individuals with five or more LTCs exhibited about 10% lower muscle strength and 5-6% lower muscle mass, placing them at a heightened risk for adverse health outcomes, including increased all-cause and cause- specific mortality.

Conditions such as RA and T2D were associated with the most severe declines in muscle strength and mass, likely due to inflammation, inactivity, and altered body composition. Interventions like resistance exercise and physical activity could mitigate these effects, improve functional outcomes, and reduce cardiovascular disease risk, given the predictive value of muscle strength for cardiovascular events (32). The findings advocate for targeted healthcare resources to benefit individuals with high multimorbidity or severe muscle impairment.

In conclusion, the study’s findings highlight a clear inverse relationship between the LTC count and both muscle strength and FFM regardless of measurements. Participants with multimorbidity, particularly those with five or more LTCs, exhibited significantly weaker muscle strength and lower FFM compared to those without LTCs. The most affected groups were individuals with type 2 diabetes, rheumatoid arthritis, gout, and coronary heart disease. Overall, the results emphasize the need for targeted interventions to maintain muscle mass and strength in individuals with LTCs, especially in those with multimorbidity.

## DATA AVAILABILITY

The data that supports the findings of this study are available upon reasonable request from the corresponding author (CC-M).

## ACKNOWLEDGEMENTS

We are grateful to the UK Biobank participants. This research has been conducted using the UK Biobank resource under application number 71932. IRG is supported by a Sara Borrell contract from Instituto de Salud Carlos III (CD23/00236).

## AUTHOR CONTRIBUTIONS

Marion Guerrero-Wyss (MGW), Frederick K Ho (FH), Bhautesh D Jani (BJ), Stuart R Gray (SG), and Carlos A Celis-Morales (CCM) designed the study. MGW and CCM analysed the data. All authors wrote the paper and have read and agreed to the published version of the paper.

## ETHICAL APPROVAL

The study was approved by the Northwest Multicentre Research Ethics Committee. This study is part of the UK Biobank project 7155 (NHS National Research Ethics Service16/NW/0274). The study protocol is available online (http://www.ukbiobank.ac.uk/).

## ADDITIONAL INFORMATION

Correspondence and requests for materials should be addressed to Carlos Celis-Morales.

