## Supplementary figures and images for "Muscle strength and muscle mass in people with long term conditions and multimorbidity: A cross-sectional study of UK Biobank participants"

### Supplemental Figure 1

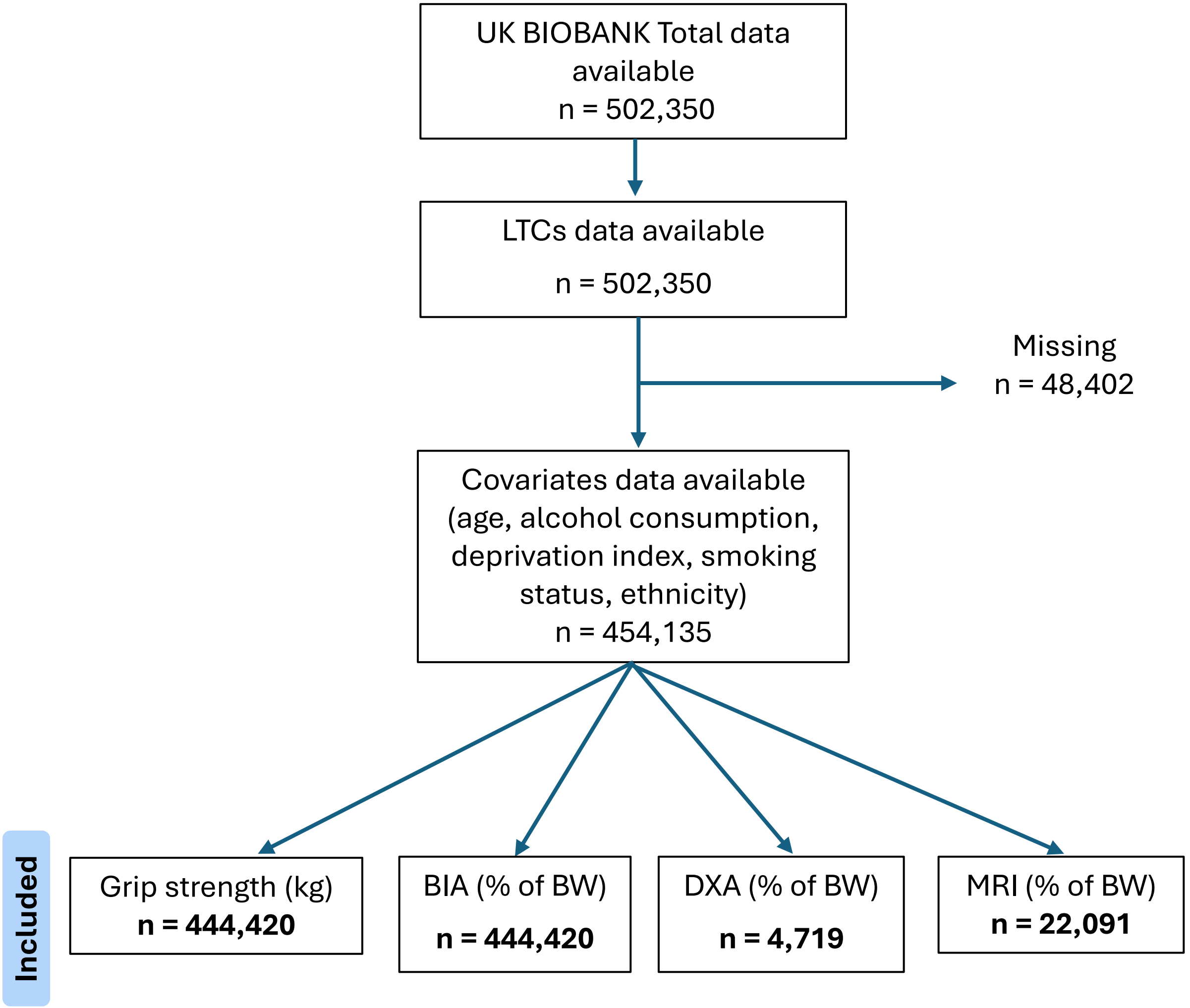

### Supplemental Figure 2

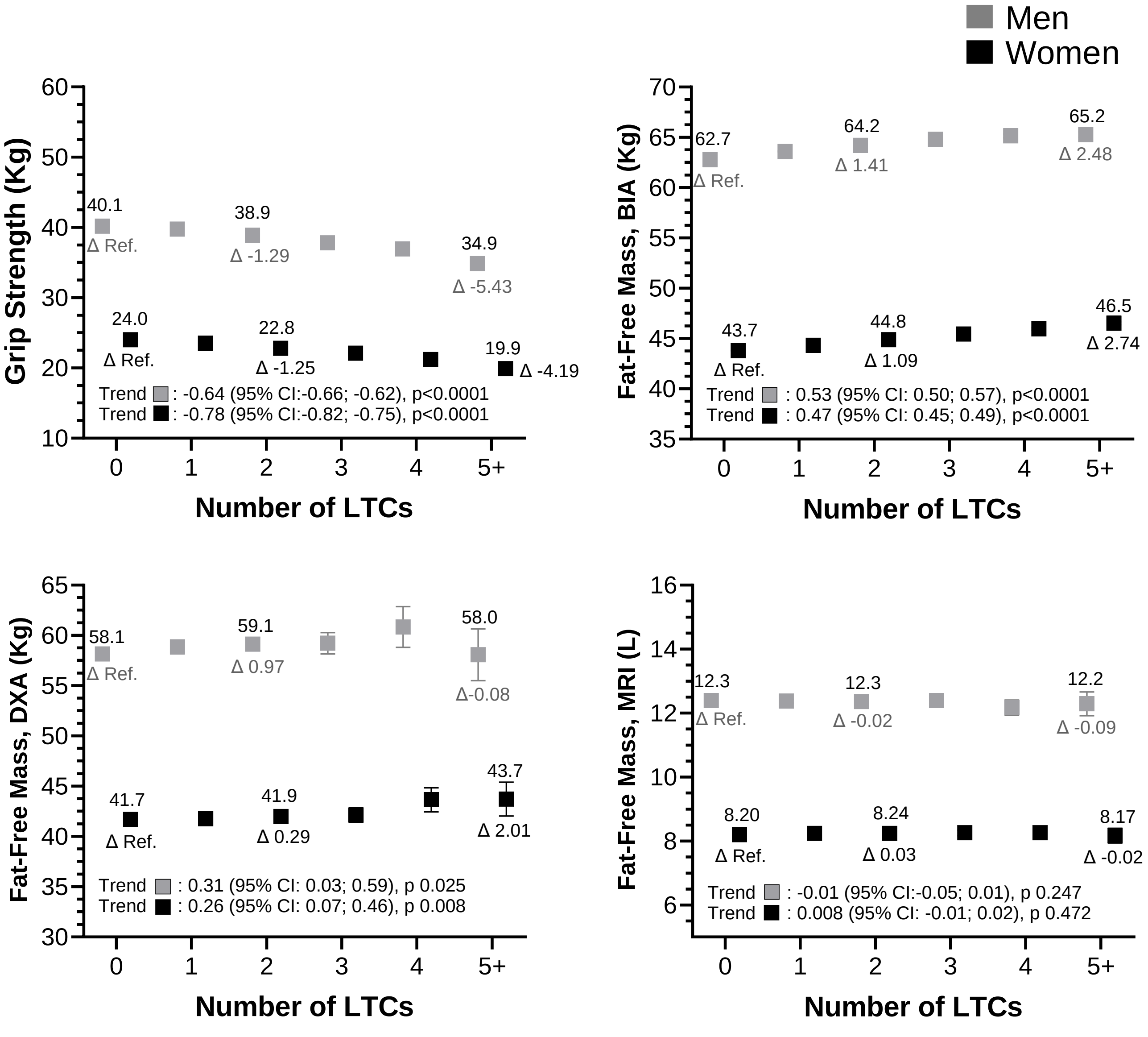
