## Supplemental Figure 3 for "Muscle strength and muscle mass in people with long term conditions and multimorbidity: A cross-sectional study of UK Biobank participants"

**Women**

| | Grip (%) | BIA FFM (%) | DXA FFM (%) | MRI FFM (%) |
|---|---|---|---|---|
| **Grip (%)** | 1 | | | |
| **BIA FFM (%)** | 0.589 | 1 | | |
| **DXA FFM (%)** | 0.590 | 0.878 | 1 | |
| **MRI FFM (%)** | 0.578 | 0.760 | 0.902 | 1 |

**Men**

| | Grip (%) | BIA FFM (%) | DXA FFM (%) | MRI FFM (%) |
|---|---|---|---|---|
| Grip (%) | 1 | | | |
| BIA FFM (%) | 0.534 | 1 | | |
| DXA FFM (%) | 0.568 | 0.837 | 1 | |
| MRI FFM (%) | 0.476 | 0.688 | 0.879 | 1 |
